# A Pilot Randomized Trial of Home-Based, Video-Supervised Exercise on Muscle Metabolism and Physical Endurance in Chronic Kidney Disease

**DOI:** 10.1101/2025.05.27.25328460

**Authors:** Gwénaëlle Begue, Armin Ahmadi, Christopher MT Hayden, Alec Foster, Usman Rehman, Jennifer E Norman, Chenoa Vargas, Brian J Bennett, Craig McDonald, T. Alp Ikizler, Hiba Hamdan, Lucas Smith, Tae Youn Kim, Thomas Jue, Jorge Gamboa, Baback Roshanravan

## Abstract

**Background:** Muscle impairment in chronic kidney disease (CKD) contributes to decreased physical performance, frailty, and higher mortality risk. Regular exercise improves muscle function in CKD. This pilot randomized controlled study evaluated the efficacy of a home-based, video-supervised exercise program on muscle function and physical endurance in CKD.

**Methods:** Sedentary adults (n=32) with moderate-to-severe nondialysis CKD (eGFR <60 mL/min/1.73m^2^) were randomized to 12 weeks of moderately intense home-based, video-supervised exercise or usual care. Co-primary outcomes included *in-vivo* muscle mitochondrial bioenergetics (rate of phosphocreatine [PCr] recovery, k_PCr_) using phosphorus-31 (^31^P) magnetic resonance spectroscopy and work efficiency using graded cycle exercise testing. Secondary outcomes included 6-minute walk distance test (6MWD), total work, and peak oxygen consumption (VO_2_peak). Other outcomes were body composition measures and plasma cytokines. Linear mixed models estimated between-group differences.

**Results:** Participants included 23 exercisers (EX) and nine in usual care (UC), with mean (*SD*) ages of 62.6 (10.8) and 67.2 (8.2) years, and eGFRs of 35.0 (12.6) and 32.3 (12) mL/min/1.73m^2^, respectively. No serious adverse events occurred; 90.5% of EX completed ≥75% of sessions. Compared to UC, EX resulted significantly increased *in-vivo* muscle mitochondrial bioenergetics (0.20min^−1^, 95%CI [0.05,0.35], *P*=0.01), total work (5.03kJ, 95%CI [1.25,8.80], *P*=0.007), and 6MWD (39.1m, 95%CI [7.1,71.1], *P*=0.014). EX preserved fat-free mass (2.23kg, 95%CI [0.46, 4.0], *P*=0.011) and marginally decreased fat mass (-2.05kg, 95%CI [-4.5, 0.37], *P*=0.087) compared to UC. IL-8 concentration differed most between EX vs. UC (effect size -1.23, 95%CI [-0.67, -0.02], *P*=0.016). Differences in IL-6, TNF-α, IL-1β, IL-10, VO_2_peak and work efficiency were non-significant between groups.

**Conclusions:** Among adults with stage 3-5 CKD, 12-weeks of moderately intense home-based video-supervised, personalized exercise is feasible and improves muscle oxidative capacity and physical endurance. By addressing common barriers to exercise, such exercise protocols could help mitigate the functional decline and frailty associated with CKD.

**Key Points:**

- A home-based video-supervised exercise program was feasible with a high level of adherence in nondialysis chronic kidney disease (CKD).
- A 12-week moderately intense home-based exercise program improved muscle mitochondria oxidative capacity and physical endurance in nondialysis CKD.
- Addressing common barriers to exercise could help mitigate the functional decline and frailty associated with CKD.

## Introduction

Chronic kidney disease (CKD), which affects 14% of the global population, leads to reduced quality of life and increased cardiovascular risks^1^. Frailty is also common in people with CKD due in part to muscle impairment caused by sedentarism and lack of physical activity^2^. Over time, the loss of muscle function known as sarcopenia contributes to decreased functional mobility, increased fatigue, decreased independence, and ultimately increased the risk of dialysis initiation, mortality and negatively impacts patients’ quality of life^3–5^. Dialysis initiation contributes to a higher risk of disability^6^. Therefore, it is important to develop disease-modifying strategies to slow the functional decline in CKD before dialysis initiation.

Muscle impairment is the final common pathway of the pathophysiology of CKD leading to functional decline^7^. Muscle mitochondrial dysfunction is mechanistically linked to low physical endurance, a key component of loss of functional mobility in people with CKD^8,9^ associated with greater risk of mortality^3^. Reduced muscle mitochondrial function in CKD has been associated with increased chronic inflammation, oxidative stress, and is also directly linked to decreased muscle performance and exercise tolerance^8,10,11^. Decreased mitochondrial bioenergetics is associated to poor physical performance on the 6-minute walk test of mobility endurance^12,13^.

Exercise training is a promising non-pharmaceutical intervention strategy to slow disease progression and improve quality of life and mobility endurance in CKD^14–16^. In the general population, exercise is an important therapeutic strategy that reduces inflammation, oxidative stress, and improves muscle mitochondrial metabolism^14–16^. Supervised home-based exercise programs may help increase accessibility and decrease practical barriers to exercise in CKD patients^15^. There is a lack of randomized controlled trials of home-based, personalized exercise training testing improvements in muscle metabolic health, muscle function, and physical endurance in nondialysis CKD^17,18^.

The present study is a pilot randomized controlled trial (NCT02923063) testing the feasibility, safety, and efficacy of a 12-week home-based, video-supervised, and personalized exercise program compared to usual care in moderate-severe (i.e., stage 3-5), sedentary, and nondialysis CKD to improve muscle metabolic health, muscle performance, and physical endurance. We assessed the efficacy of the exercise intervention on the primary endpoints of *in-vivo* muscle mitochondrial oxidative capacity and muscle performance by submaximal work efficiency using cycle ergometer graded exercise testing (GXT). The secondary endpoints of physical endurance included total muscle work and performance at the 6-minute walk test (6MWT). Feasibility and safety endpoints included participant adherence and adverse events related to the exercise program.

## Methods

### Study population and design

Exercise Study Testing Enhanced Energetics of Muscle Mitochondria-Video Integrated Delivery of Activity (ESTEEM-VIDA) is a pilot randomized-controlled trial (NCT02923063) including moderate-severe nondialysis individuals with CKD (eGFR<60ml/min per 1.73m^2^). Participants were recruited by clinical research coordinators who approached eligible patients seen in nephrology clinics within the University of California Davis Health system throughout the greater Sacramento area (California, USA). The clinical trial was approved by the university institutional review board (ID 1343904-21) and conducted from January 2020 to March 2024 including during the COVID-19 pandemic (2020-2022). A schematic of the study design and study endpoints is presented in **Figure 1**, and the CONSORT diagram is presented in **Figure 2**.

**Figure 1.**
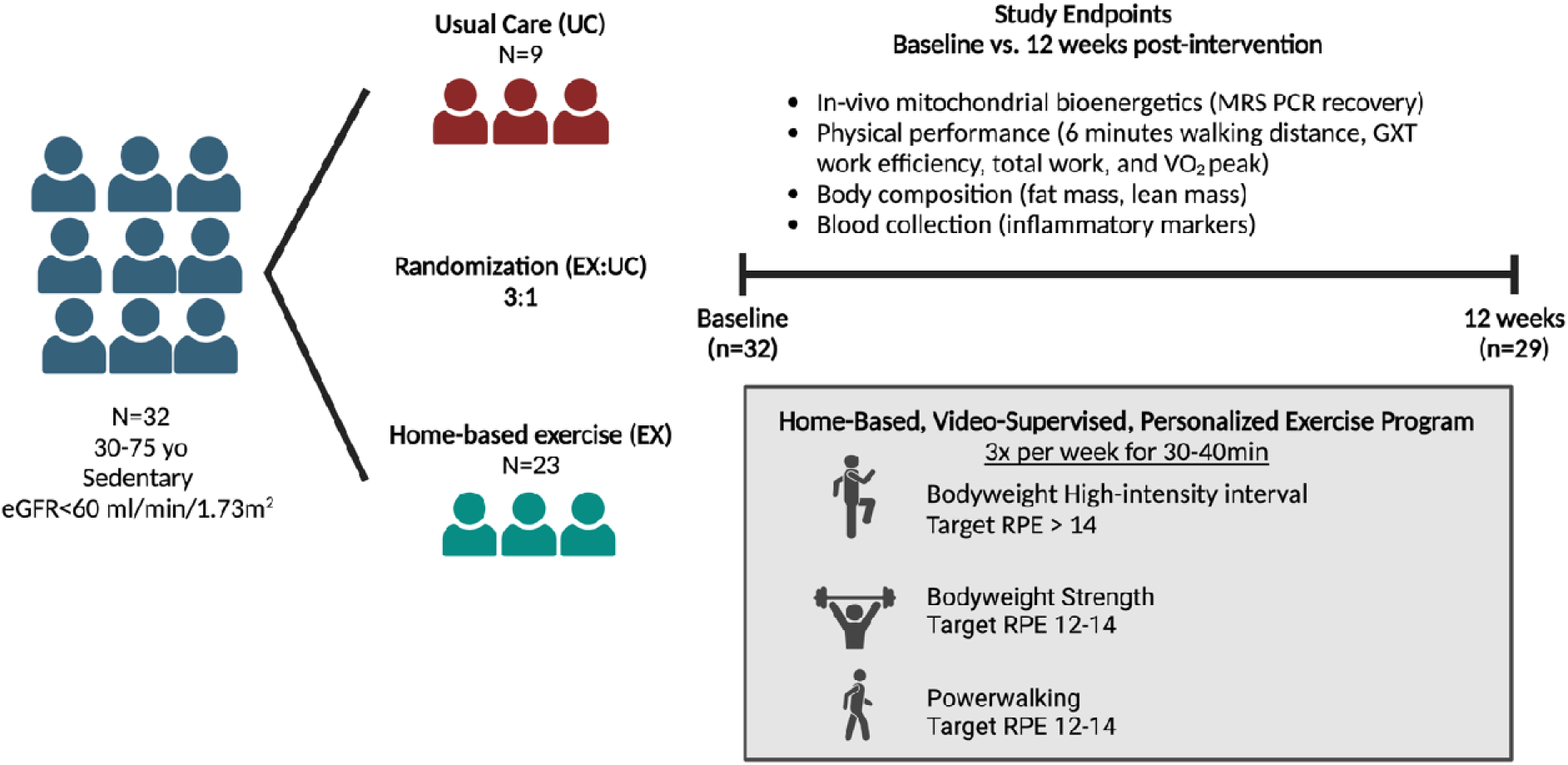
Overview of study design.

**Figure 2.**
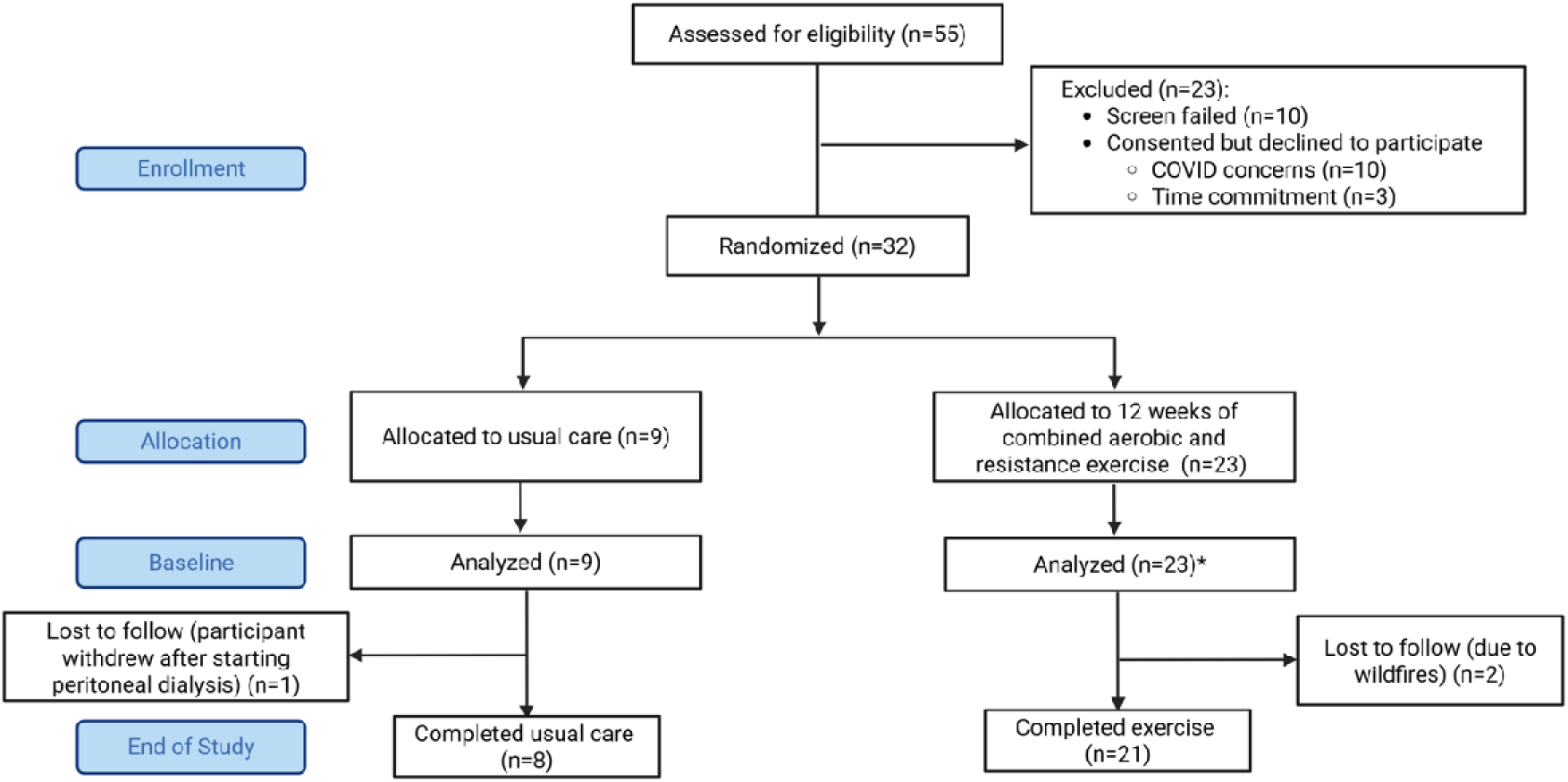
CONSORT diagram for ESTEEM-VIDA.

Individuals with an eGFR of <60ml/min per 1.73m^2^, age of 30-75years, and sedentary as defined by the Molecular Transducers of Physical Activity Consortium^19^ were eligible to participate. Main exclusion criteria included: uncontrolled diabetes (HgbA1c >8.5), drugs or supplements that are known to alter mitochondrial function, contraindications to participate in vigorous exercise, uncontrolled hypertension (systolic >170, diastolic >100mmHg), and implants that prohibit magnetic resonance imaging (MRI) measurements **(Supplemental Table 1)**.

Pre-screening and consenting were conducted remotely via phone or videoconference. Participants were randomly assigned to the usual care (UC) or exercise (EX) group. A randomization table stratified by sex and diabetes was created with a 3:1 allocation ratio (EX:UC) using R package blockrand (version 1.5). The allocation table was then implemented in REDCap where the randomization sequence was concealed from the investigators and study coordinator. After determining eligibility of each participant, the study coordinator randomized the participant in REDCap (**Figure 2**). The remote baseline visit included a detailed assessment of demographics, smoking, medical history, and medication inventory including over-the-counter medications.

The pre- and post-intervention visits involved fasting blood collection followed by *in-vivo* muscle mitochondrial bioenergetics measurement, anthropometrics, vital signs, body composition analysis, medical history and exam performed by the study clinician, measurement of oxygen consumption at rest and during exercise (**Figure 1**). Testing was conducted in 1 to 2 visits depending on participant preference and whether the visits fell during COVID-19 restrictions.

### Study Endpoints

The primary endpoints of muscle function were 1) *in-vivo* muscle mitochondria oxidative capacity assessed via measuring the rate of phosphocreatine (PCr) recovery (k_PCr_) using ^31^P-MRS following a brief exercise, and 2) submaximal work efficiency calculated as gross efficiency using oxygen consumption (VO_2_) and respiratory exchange ratio (RER, i.e., VCO_2_/VO_2_) measured during a GXT. The secondary endpoints of physical endurance and cardiorespiratory fitness (CRF, i.e., aerobic capacity) were total work and peak oxygen consumption (VO_2_peak) measured during the GXT as well as the 6MWT distance. Feasibility and safety endpoints included participant adherence and adverse events related to the exercise program. Other endpoints included body composition measures and plasma cytokine levels.

### Home-based video-supervised exercise program

The exercise (EX) group followed a 12-week moderately intense home-based, personalized, and video-supervised exercise program. Participants exercised 3 times a week starting at 30min per session and increasing by 5min every 4 weeks up to 40min at the end of the 12 weeks (**Figure 1**). The 3 weekly sessions included: one session of 1) bodyweight high-intensity interval training (HIIT), 2) strength training, and 3) power walking. Bodyweight HIIT sessions targeted a rate of perceived exertion (RPE) of 15 or above (Borg scale, range 6-20)^20^. Strength training sessions included bodyweight to low free weight and resistance band movements targeting 10-20 repetitions per exercise. Both strength and power walking sessions targeted an RPE between 12 to 14 (i.e., moderate intensity). The exercise program was personalized to each participant’s physical capabilities and mobility determined using the Selective Functional Movement Assessment (SFMA)^21^ administered online and laboratory pre-intervention physical performance. Participants completed one supervised week of Zoom-based exercise sessions (Zoom Video Communication Inc.) with a trainer, followed by one self-directed week using pre-recorded personalized videos. Trainers recorded these videos based on previous sessions to ensure familiarity. Videos were uploaded to Vimeo®, and personalized instructions were sent via email. On non-exercise days, participants were encouraged to walk at least 30min at a leisure pace.

Participants wore heart rate monitors (Polar H10) for every exercise session, synchronized with the *Polar Flow* cloud-based platform for monitoring adherence remotely. Exercise adherence was defined as ≥75% of the 36 total sessions over 12 weeks while safety was measured using the number of adverse events related to the exercise program. Both groups received a Go4Life “Workout to go” sample exercise routine created by the National Institutes on Aging (NIA)^22^. All participants were also given a brochure containing information on dietary recommendations as prescribed by the National Kidney Disease Education Program^23^, reviewed with the study physician during the pre-intervention visit.

### 6-minute walk test (6MWT)

The 6MWT distance (6MWD) was used as a marker of physical endurance and functional mobility. Walking economy (i.e., energy cost of walking) was calculated as relative VO_2_ (ml/kg/min) divided by walking speed (m/min) (**Supplemental Methods**).

### Graded cycle exercise testing (GXT)

Physical endurance was also assessed as total work performed on a recumbent cycle ergometer (Ergobike 600k, Cosmed) using a GXT (**Supplemental Methods**). Peak oxygen consumption (VO_2_peak) was measured to assess CRF. Work (kilojoules, kJ) performed at each stage was calculated as (watts*seconds)/1000 with total work determined by summing stage values. Gross work efficiency (%) was calculated at a submaximal workload of 35watts (W) by dividing work accomplished per minute (kcal.min^-1^) by energy expended per minute (kcal.min^-1^). Energy expenditure was calculated from VO_2_ (l/min) and RER (i.e., fuel utilized) using the non-protein RER energy equivalent table^24,25^. This workload was completed by all study participants and considered a submaximal workload according to participants peak work rate (range: 60-185W).

### In-vivo muscle oxidative capacity (^31^P-MRS)

To investigate *in-vivo* muscle oxidative capacity changes in response to exercise training, the rate of PCr recovery (k_PCr_) was measured using phosphorus-magnetic resonance spectroscopy (^31^P- MRS) after exercise (42seconds of knee extension) targeting a 20-30% PCr drop from the control level and minimal intramuscular pH change (pH>6.8). k_PCr_ can assess muscle mitochondrial bioenergetics in a broad range of subjects, including those with CKD^12^. ^31^P-MRS assessment and k_PCr_ calculations were conducted as previously described^26^ with specifics included in **Supplemental Methods**. The ^31^P-MRS data analyst was blinded to the group assignment of participants. The magnetic resonance imaging (MRI) scanner underwent an upgrade requiring unanticipated repairs and additional validation of ^31^P- MRS protocols from July 2022 to April 2023. Baseline ^31^P-MRS data were not obtained on two participants randomized to exercise but were obtained at the end of study and included in statistical analysis.

### Body Composition and Resting Energy Expenditure

Body composition was measured using bioimpedance spectroscopy (Impedimed SFB7) after participants rested for 3-5min. Resting energy expenditure (REE) was measured over 15min using a breath-by-breath metabolic analyzer system (Cosmed K5). Body composition and REE were collected after 8-12hr fast (**Supplemental Methods**).

### Plasma Cytokines

Plasma cytokines (IL-6, IL-8, TNF-α, IL-1β, IL-10) were assessed in duplicates (pg/ml) in a single multiplex electroluminescence assay (MESO QuickPlex SQ 120MM) using the proinflammatory panel 1 V-Plex kit (K15049D, Meso Scale Discovery, Rockville, MD, USA) and following the manufacturer instructions (**Supplemental Methods**).

### Statistical Analysis Plan

Linear mixed models (LMMs) were chosen to handle missing and unbalanced data related to participants lost to follow-up and MRI scanner upgrade. LMMs were used to estimate between-group differences in physical performance outcomes, body composition measures, REE, and *in-vivo* muscle oxidative capacity using the interaction of treatment (UC vs. EX) and time (pre- vs. post-intervention) with random intercepts (individual-specific baselines) and random slopes (individual-specific responses) with robust Huber-White standard errors. Within-group differences were estimated using LMMs with time (pre- vs. post-intervention) as a fixed effect and random intercepts.

#### Cytokine analysis

All inflammatory markers were checked for normality via the Shapiro test. Log-transformation was used to enhance data normality. LMMs using log-transformed inflammatory markers were used to assess within- and between-group differences. To quantify the impact of 12 weeks of exercise the effect size using standardized mean difference was used. The effect size of 0.2, 0.5, and 0.8 were, respectively, considered small, medium, and large.

#### Power calculation

The sample size was determined using the *in-vivo* leg muscle mitochondrial capacity (ATPmax) measurements from the Chronic Kidney Disease Muscle Mitochondrial ENergetics and Dysfunction (CKD-MEND) study- A cross-sectional study of 53 participants with moderate-severe CKD (mean eGFR of 33ml/min per 1.73 m^2^) and 24 healthy controls (mean eGFR of 102ml/min per 1.73 m^2^)^12^. The rate of PCr recovery (k_PCr_) was calculated from leg muscle ATPmax yielding an estimated mean of 1.37min^-1^ and *SD* of 0.36min ^-1^ among those with CKD. This difference of 0.36min-^1^ corresponded to a difference of 50meters in a 6-minute walk test within CKD. We estimated sample sizes of 42 and 32 to yield 80% or 70% power, respectively, allowing detection of a minimum difference of 0.36min^-1^ between EX and UC with an allocation ratio of 3:1 exercise to usual care at a two-sided 0.05 significance level. Unfortunately, due to delays related to COVID-19 restrictions and prolonged MRI shutdown for upgrades and repairs, the clinical trial recruited 32 participants and was terminated before achieving the original targeted recruitment of 45.

A *P-*value of less than 0.05 was considered significant in this study. Figures were generated using GraphPad Prism 10.3. Analyses were conducted in R (R Core Team) version 4.2.2 and Stata version 17.0 (StataCorp, College Station, TX).

## Results

### Participant baseline characteristics

A total of 32 participants were enrolled, 3 were lost to follow-up; leaving 29 who completed the study (**Figure 2**). Baseline characteristics of the 32 participants in this study can be found in **Table 1**.

**Table 1.** Participant characteristics of analytic population at baseline (n=32)

| Characteristics | Usual care | Exercisers |
| --- | --- | --- |
| Number | 9 | 23 |
| Demographics |  |  |
| Age, mean (*SD*) | 67.2 (8.2) | 62.6 (10.8) |
| Female, No. (%) | 7 (78) | 11 (48) |
| Race, No. (%) |  |  |
| Asian/Pacific Islander | 1 (11) | 3 (13) |
| Black | 4 (44) | 2 (9) |
| White | 4 (44) | 18 (78) |
| Medical history and lifestyle, No. (%) |  |  |
| Diabetes, No. (%) | 4 (44) | 12 (52) |
| Six-minute walking distance, (m) | 483 (77) | 452 (92) |
| VO<sub>2</sub> peak, (ml/kg/min) | 16.3 (4.1) | 16.1 (3.2) |
| Medication use, No. (%) |  |  |
| ACE Inhibitors/ARB | 6 (67) | 15 (65) |
| GLP-1 agonist | 1 (11) | 6 (26) |
| SGLT-2 inhibitor | 2 (22) | 2 (5) |
| Diuretics | 4 (44) | 6 (26) |
| β-Blockers | 4 (44) | 8 (35) |
| Calcium-channel blockers | 3 (33) | 12 (52) |
| Metformin | 2 (22) | 6 (26) |
| Statins | 3 (33) | 12 (52) |
| Physical characteristics, mean (*SD*) |  |  |
| BMI (kg/m<sup>2</sup>) | 30.7 (5.1) | 33.2 (6.7) |
| Body weight (kg) | 80.3 (14.9) | 93.6 (20.7) |
| Fat-free mass (kg) | 55.8 (10.3) | 60.8 (15.0) |
| Percent fat-free mass (%) | 70.3 (11.7) | 65.1 (8.5) |
| Fat mass (kg) | 28.5 (9.3) | 33.3 (11.6) |
| Percent fat mass (%) | 36.5 (13.9) | 35.5 (9.6) |
| Laboratory data |  |  |
| Serum creatinine (mg/dl), median (IQR) | 1.6 (1.4 to 2.0) | 1.6 (1.4 to 2.6) |
| eGFR (mL/min/1.73 m<sup>2</sup>), mean (*SD*) | 32.3 (12) | 35.0 (12.6) |
| Hemoglobin (g/dL), mean (*SD*) | 12.6 (2.3) | 13.2 (1.7) |

Data are means (*SDs*) or medians (*IQR*) for continuous variables, and number (%) for categorical variables.
Abbreviations: *SD*, standard deviation; *IQR*, interquartile range; No, number; HAP, human activity profile;
ACEi/ARB, angiotensin converting enzyme inhibitors/angiotensin-receptor blockers; GLP-1, glucagon-Like
Peptide-1; SGLT-2, Sodium-Glucose Cotransporter-2; eGFR, estimated glomerular filtration rate.

Participants in the UC had a mean ± *SD* age of 67.2 ± 8.2years and mean eGFR of 32 ± 2mL/min/1.73m^2^. Those in the EX-group had a mean age of 62.6 ± 10.8years and mean eGFR of 35 ± 13mL/min/1.73m^2^. UC had more females than EX (78% vs. 48%) and participants with a diagnosis of diabetes were 44% and 52% for UC and EX, respectively. Baseline peak VO_2_ were similar for UC and EX and considered below 50^th^ percentile for age and sex^27^.

### Twelve weeks of moderately intense home-based video-supervised exercise was feasible with high participant adherence

Out of 23 participants randomized to EX, 21 (91%) completed the 12-weeks exercise program with two lost to follow-up (**Figure 2**). The average adherence was 90.5% with 32 of 36 sessions (range: 14-36) completed. Only two (9.5%) exercisers reported less than 75% adherence. Exercisers reported an average RPE of 14.3 ± 1.3 compared to targeted RPE of above 14 for HIIT session, and RPEs of 13.9 ± 1.8 for strength and powerwalking sessions compared to targeted RPE of 12-14. A higher number of adverse events were noted in the EX-group compared to UC; however, none were thought to be related to the exercise protocol (**Supplemental Table 2**).

### Twelve weeks of moderately intense home-based exercise improved in-vivo muscle oxidative capacity compared to usual care

The primary endpoint*, in-vivo* muscle oxidative capacity (k_PCr_), is presented in **Table 2** and **Figure 3**. Compared to UC, the EX-group significantly increased their muscle oxidative capacity. Complete case analysis (n=28) yielded similar results with the EX-group (n=20) improving muscle oxidative capacity by an estimated 0.18min^-1^ (95%CI [0.04, 0.33], *P*=0.014) compared to UC (n=8).

**Figure 3.**
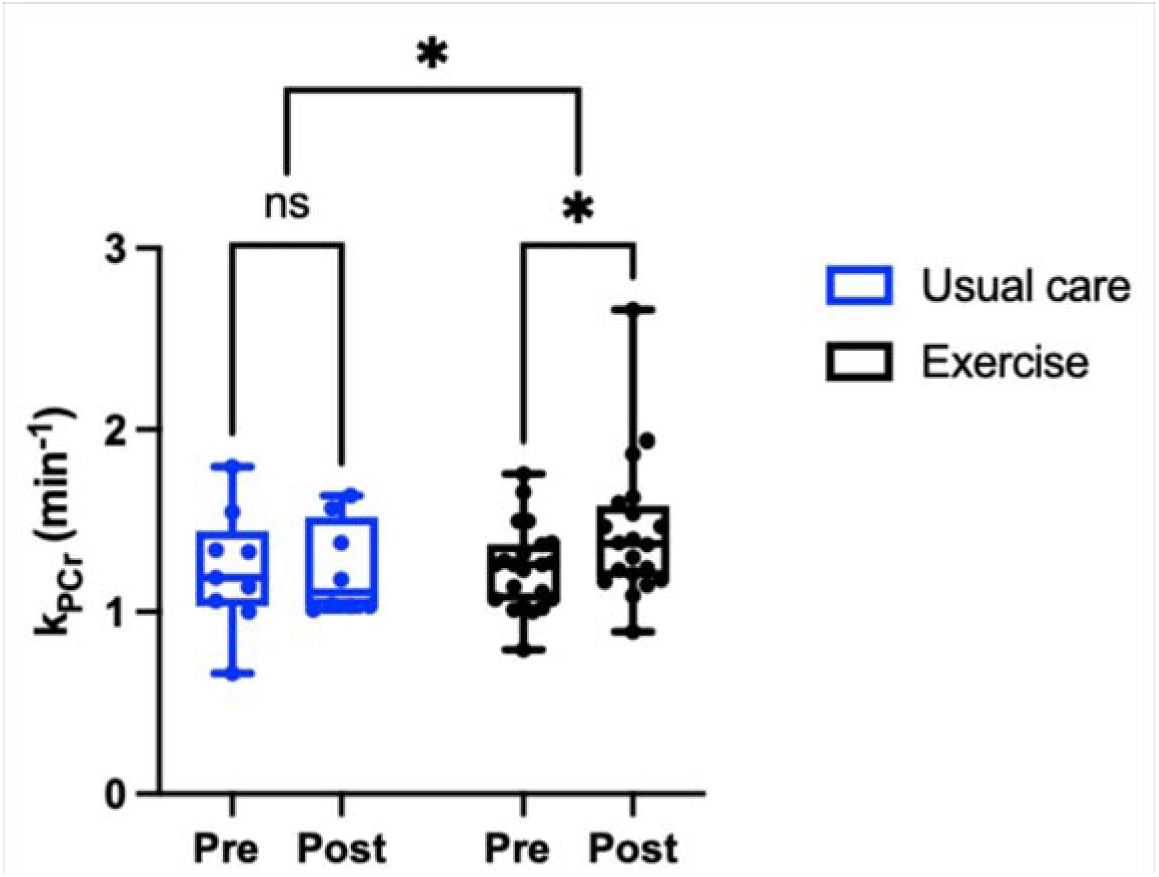
Vastus lateralis muscle phosphocreatine recovery (k_PCr_) after a brief dynamic exercise (n=32).

**Table 2.** Summary of primary and secondary endpoints comparing usual care and exercise over the 12-week intervention. Linear mixed effects modeling was used to estimate the effect of exercise compared to usual care. Mean differences, 95%CIs, and *P*-values are shown. The values in bold represent *P* <0.05.

| Endpoint | Usual Care |  | Exercise |  | Within group differences |  | Between group differences |  |
| --- | --- | --- | --- | --- | --- | --- | --- | --- |
|  | Baseline mean (*SD*) | 12 weeks mean (*SD*) | Baseline mean (*SD*) | 12 weeks mean (*SD*) | Usual care Beta (95% CI) | Exercise Beta (95% CI) | Exercise-usual care Beta (95% CI) | Treatment *P*-value |
| **<sup>31</sup>P-MRS** |  |  |  |  |  |  |  |  |
| k<sub>PCr</sub> (min<sup>-1</sup>) | 1.23 (0.33) | 1.24 (0.23) | 1.24 (0.26) | 1.44 (0.38) | -0.06 (-0.16, 0.04) | **0.15 (0.02, 0.28)** | **0.20 (0.05, 0.35)** | 0.01 |
| **Graded Exercise Test** |  |  |  |  |  |  |  |  |
| Work Efficiency at 35W (%) | 14.3 (2.2) | 13.4 (2.8) | 14.0 (2.3) | 13.0 (2.9) | -0.28 (-1.88, 1.31) | -0.21 (-1.22, 0.78) | 0.08 (-1.81, 1.98) | 0.93 |
| Total Work (kJ) | 27 (17.8) | 29.3 (15.4) | 22.4 (15.4) | 32.4 (18) | -1.55 (-3.99, 0.88) | **3.56 (0.72, 6.45)** | **5.03 (1.25, 8.80)** | 0.007 |
| VO<sub>2</sub> Peak (ml/kg/min) | 16.3 (4.1) | 16.1 (3.2) | 17.3 (3.7) | 17.9 (3.8) | 1.17 (-1.77, 4.11) | **1.84 (0.78, 2.9)** | 0.59 (-2.58, 3.77) | 0.70 |
| VO<sub>2</sub> Peak (ml/kg FFM/min) | 24 (4) | 24.7 (4.4) | 23 (10.8) | 26.5 (5.2) | -1.04 (-8.4, 6.3) | 1.5 (-0.38, 3.5) | 2.6 (-5, 10.2) | 0.51 |
| VO<sub>2</sub> Peak (l/min) | 1.35 (0.38) | 1.5 (0.41) | 1.33 (0.36) | 1.67 (0.52) | 0.016 (-0.19, 0.23) | **0.17 (0.07, 0.27)** | 0.133 (-0.10, 0.37) | 0.24 |
| **6-minute Walk Test** |  |  |  |  |  |  |  |  |
| Distance (m) | 483 (77) | 452 (92) | 465 (61) | 481 (75) | -10.3 (-31.5, 10.9) | **28 (3.7, 52)** | **39.1 (7.1, 71.1)** | 0.014 |

### Twelve weeks of moderately intense home-based exercise improved physical endurance, but not cardiorespiratory fitness and metabolic efficiency compared to usual care

Primary and secondary endpoints of work efficiency, physical endurance, and CRF are shown in **Figure 4** and **Table 2**. Work efficiency (%) measured at submaximal workload of 35W did not change within- or between-groups. This is supported by no changes in submaximal absolute VO_2_ and fuel utilization (RER) at 35W within or between groups (**Supplemental Table 3**). Compared to UC total work increased with EX with an estimated between-group difference of 5.03kJ (95%CI [1.25,8.80], *P*=0.007) paralleling increases in total GXT duration (**Supplemental Table 3**). Exercisers increased their 6MWD by 28m (95%CI [3.7, 52], *P*=0.02) with a difference between groups of 39m (95%CI [7.8, 70.3]; *P*=0.01). Absolute and relative to body weight VO_2_ peak significantly increased in EX without any change in VO_2_peak relative to fat-free mass, but between-group changes were not statistically significant.

**Figure 4.**
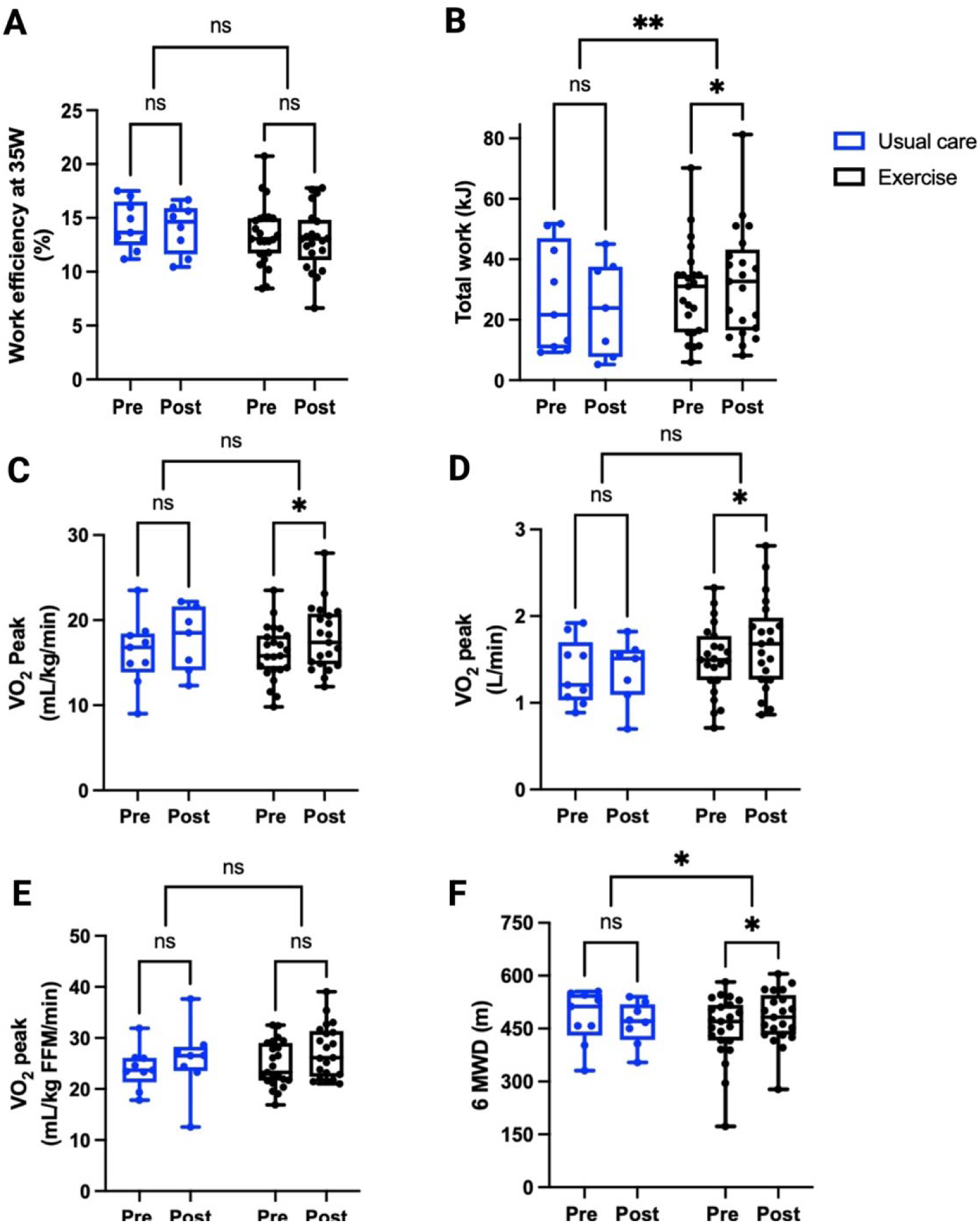
Changes in physical performance, cardiorespiratory fitness, and efficiency comparing usual care vs exercise group (n=32).

Exploratory endpoints of economy and efficiency are shown in **Supplemental Table 3**.

Exploratory changes in physical performance, CRF, and efficiency endpoints among EX stratified by sex and diabetes status are shown in **Supplemental Figures 1** and **2**.

### Twelve weeks of home-based exercise favorably remodeled body composition towards lower fat mass and maintained fat-free mass compared to usual care

Body composition measures are presented in **Table 3**. UC significantly lost fat-free mass while EX maintained their fat-free mass leading to a between-group differences of 2.23kg (95%CI [0.46,4.0], *P*=0.01). The percentage of fat-free mass relative to body weight increased by an average of 1.7% within EX with a between group differences of 2.6% (95%CI [0.07,5.4], *P*=0.04) compared to UC. Body composition remodeling did not significantly affect resting energy expenditure within EX and UC.

**Table 3.** Changes in body composition endpoints comparing the usual care and the exercise group (n=32). Linear mixed effects modeling was used to estimate the effect of exercise compared to usual care. Mean differences, 95%CIs, and *P*-values are shown. The values in bold represent *P* <0.05.

| Endpoint | Usual Care |  | Exercise |  | Within group differences |  | Between group differences |  |
| --- | --- | --- | --- | --- | --- | --- | --- | --- |
|  | Baseline mean (*SD*) | 12 weeks mean (*SD*) | Baseline mean (*SD*) | 12 weeks mean (*SD*) | Usual care Beta (95% CI) | Exercise Beta (95% CI) | Exercise-usual care Beta (95% CI) | Treatment *P*-value |
| Body weight, (kg) | 80.3 (14.9) | 79.2 (14.7) | 93.6 (20.7) | 92.6 (20.7) | -1.12 (-2.75, 0.51) | **-1.10 (-2.13, -0.08)** | 0.02 (-1.90, 1.94) | 0.98 |
| Fat-free mass (kg) | 55.8 (10.3) | 54.5 (11.2) | 60.8 (15.0) | 60.8 (14.7) | **-1.36 (-2.57, -0.16)** | 0.85 (-0.43, 2.14) | **2.23 (0.46, 4.0)** | 0.011 |
| Percent fat-free mass (%) | 70.3 (11.7) | 69.5 (12.9) | 65.1 (8.5) | 65.8 (7) | -0.85 (-3.2, 1.5) | **1.7 (0.3, 3.0)** | **2.55 (0.07, 5.4)** | 0.044 |
| Fat mass (kg) | 28.5 (9.2) | 28.9 (8.1) | 33.3 (11.6) | 32.3 (10.6) | 0.46 (-1.57, 2.49) | **-1.6 (-2.9, -0.28)** | -2.05 (-4.5, 0.37) | 0.087 |
| Percent fat mass (%) | 36.5 (13.9) | 37.9 (14.2) | 35.5 (9.6) | 35.4 (8.7) | 1.47 (-0.7, 3.6) | **-1.24 (-2.4, -0.09)** | **-2.72 (-5.2, -0.29)** | 0.04 |
| Resting Energy Expenditure (kcal/day) | 1511(164) | 1370 (286) | 1628 (344) | 1645 (458) | -147 (-404, 111) | 29 (-86, 145) | 204 (-83, 490) | 0.19 |

### Home-based exercise and cytokine response

Cytokine levels are presented in **Table 4**. IL-8 was the only inflammatory cytokine that differed between EX and UC. IL-8 increased in UC while IL-8 did not change within EX. IL-8 between group differences was a large effect size of -1.23 (95%CI [-0.67, -0.02], *P*=0.016). There were no significant within- or between-group differences on other measured cytokines. The mean coefficient of variation was 3.16 ± 5.5% across all measured cytokines.

**Table 4.** Summary of plasma cytokine endpoints (n=32). Linear mixed effects modeling was used to estimate the impact of exercise compared to usual care. Effect sizes, 95%CIs, and *P*-values are shown. The values in bold represent *P*<0.05.

| Cytokine | Usual Care |  | Exercise |  | Within group differences |  | Between group differences |  |
| --- | --- | --- | --- | --- | --- | --- | --- | --- |
|  | Baseline<br>mean (*SD*) | 12 weeks<br>mean (*SD*) | Baseline<br>mean (*SD*) | 12 weeks<br>mean (*SD*) | Usual care<br>effect size<br>(95% CI) | Exercise<br>effect size<br>(95% CI) | Exercise-usual care<br>effect size (95% CI) | Treatment<br>*P*-value |
| TNF-α<br>(pg/ml) | 5.6 (8.3) | 3.4 (3.5) | 2.3 (2.0) | 1.9 (1.4) | -0.08 (-0.41, 0.34) | -0.06 (-0.14, 0.11) | 0.07 (-0.30, 0.34) | 0.83 |
| IL-8<br>(pg/ml) | 3.9 (4.3) | 5.3 (2.8) | 4.2 (5.2) | 2.3 (1.6) | 0.80 (-0.05, 0.47) | -0.50 (-0.29, 0.03) | **-1.23 (-0.67, -0.02)** | 0.016 |
| IL-6<br>(pg/ml) | 3.3 (2) | 4 (1.4) | 3.2 (3.4) | 4.8 (7.1) | 0.53 (-0.08, 0.30) | 0.24 (-0.12, 0.28) | -0.08 (-0.38, 0.32) | 0.57 |
| IL-1β<br>(pg/ml) | 28.5 (31) | 24.5 (28) | 13.3 (11.7) | 20.5 (17.3) | -0.32 (-0.47, 0.25) | 0.43 (-0.10, 0.43) | 0.75 (-0.21, 0.81) | 0.33 |
| IL-10<br>(pg/ml) | 5.5 (3.1) | 5.7 (3.8) | 2.1 (1.4) | 2.8 (2.3) | 0.11 (-0.25, 0.20) | 0.28 (-0.07, 0.19) | 0.43 (-0.15, 0.33) | 0.71 |
| IL-6/IL-10 | 0.7 (0.5) | 2.3 (4.7) | 2.3 (3.3) | 4 (7.2) | 0.8 (-0.05, 0.47) | 0.24 (-0.58, 1.35) | -0.29 (-1.84, 1.14) | 0.64 |

## Discussion

The ESTEEM-VIDA pilot randomized-controlled study provided preliminary evidence supporting the feasibility and safety of a 12-week, home-based, video-supervised and personalized exercise program for improving muscle metabolic health and physical endurance in sedentary individuals with moderate-to-severe CKD. Despite not reaching our initial recruitment target, our findings suggest that a moderately intense home-based combined aerobic and resistance exercise program may enhance primary outcome of muscle mitochondrial oxidative capacity as well as improve secondary outcomes of physical endurance. Body composition showed a favorable response following exercise, with preserved fat-free mass and reduced fat mass compared to usual care. Across the 5 measured cytokines, IL-8 differed most between UC and EX over the 12-week period. However, no significant differences were observed for work efficiency and cardiorespiratory fitness.

Unlike prior studies, which often rely on in-person intradialytic or gym-based protocols, home-based video-supervised exercise serves to overcome critical barriers to exercise including limited access to gym facilities, economic constraints, and social distancing requirements, as experienced during the COVID-19 pandemic. High adherence rates (90.5%) and safety profile with no serious exercise-associated adverse events underscore the program’s practical applicability. Social interactions with personal trainers and the personalized approach to exercise might be contributing factors to adherence and safety of the program. Our study findings also aligned with data in individuals with heart failure, which demonstrated the equal effectiveness of home-based cardiac rehabilitation compared to center-based programs to improve exercise capacity and mortality risks^28^.

Muscle mitochondrial oxidative capacity was significantly and meaningfully improved despite our prescribed moderately intense dose of personalized training. Muscle mitochondrial dysfunction is mechanistically linked to impaired muscle performance and cardiorespiratory fitness^29^. ^31^P-MRS is the gold-standard, non-invasive technique for assessing *in-vivo* muscle oxidative capacity^30^ with a faster k_PCr_ associated with greater leg power and cardiorespiratory fitness^31^ and lower risk of future mobility decline in older adults^9^. The observed improvement in k_PCr_ with exercise coincided with clinically relevant gains in mobility endurance and muscle endurance with a39m difference in the 6MWD^32^ and a significant increase in total work output on cycle ergometry in EX versus UC. A moderately intense home-based exercise approach may serve as a practical and efficacious modality to improve metabolic health and physical endurance in persons living with CKD

Despite gains in endurance, we did not observe significant improvements in muscle work efficiency and VO_2_peak with exercise compared to usual care. The GXT protocol included 2min stages in 25W increment, which may not be ideal to effectively measure efficiency considering that it may require longer than 2min for naïve exercisers to reach steady-state oxygen consumption, a key factor for aerobic energy expenditure measurement. Furthermore, although the 35W workload was submaximal for most of our participants, it was close to maximal peak power output for a few; impacting the reliability of aerobic energy efficiency measurement in our study. Therefore, future studies should consider different GXT protocols to reliably measure work efficiency that include longer stages and smaller increases in workload.

CRF or VO_2_peak is a critical predictor of all-cause mortality and functional independence in both the general population and individuals with CKD^33,34^. Among exercisers, the relative VO_2_peak showed a modest increase. This was primarily attributed to decreased fat mass, as fat-free mass remained unchanged. Additionally, the small but significant increase in absolute VO_2_peak among exercisers suggests that the intervention influenced CRF independent of body composition changes. The lack of between-group differences in CRF could be attributed to the low dose of high-intensity interval training (HIIT), limited to once per week^35,36^. Only one previous pilot study showed that HIIT performed twice weekly for 20-30min was feasible and increased peak power output and physical functioning, but not muscle mass in dialysis-dependent CKD^37^. Future studies should test the feasibility of personalized exercise programs with higher doses of HIIT to maximize gains in physical functioning and CRF.

Chronic inflammation associated with CKD is linked to mortality across all disease stages^38,39^. While the anti-inflammatory effect of exercise training has been well-studied in various populations, several studies in nondialysis CKD have reported mixed results ^17,40^. We found IL-8 was the only cytokine that differed between exercise versus usual care over twelve weeks. This may be attributed to the small sample size and variability in cytokine responses. IL-8 is a pro-inflammatory cytokine known to recruit neutrophils to sites of inflammation and promote angiogenesis^41^. Elevated IL-8 levels have been associated with chronic inflammation and muscle wasting in older adults^42,43^. A prior large, randomized controlled trial of 12 months of moderate intensity physical activity in older adults demonstrated IL-8 to be the only cytokine responsive to the intervention^44^. Reduction in IL-8 has also been observed in randomized trials of exercise training in metabolic syndrome patients^45,46^ but data are lacking on potential IL-8 association with clinical outcomes.

### Strengths & Limitations

The strengths of our pilot home-based exercise program included its integration of remote monitoring technology and tailored approach. Another strength included the use of rigorous CRF testing and the use of innovative ^31^P-MRS to assess changes in muscle mitochondrial energetics to provide a comprehensive assessment of muscle adaptation to exercise. The primary limitation of this study is its small sample size, which was further impacted by the COVID-19 pandemic and an extended shutdown of the MRI operations delaying recruitment and leading to early study termination. The intervention’s relatively short duration and low dose of HIIT might have also limited its potential to significantly improve CRF, work efficiency, and overall muscle metabolic health.targeting recruitment of sedentary persons with CKD, the relatively select group of participants in this pilot trial may reduce generalizability to patients with more severe CKD with greater comorbid burden ^47^.

### Conclusion and Future Directions

The moderately intense home-based, video-supervised, and personalized exercise protocol addresses common barriers to exercise and can help mitigate the functional decline and frailty associated with CKD, ultimately reducing the burden on patients and healthcare systems. Such programs have the potential to enhance muscle mitochondria function, leading to a clinically meaningful increase in physical endurance. Future larger studies are needed to investigate the impact of higher doses of HIIT within home-based exercise programs to optimize cardiorespiratory fitness, muscle metabolic health, and ultimately reduce fatigue in this population.

## Data Availability

All data produced in the present work are contained in the manuscript

## Disclosures

Authors have no conflict of interest to disclose.

## Author Contributions

BR, JG, TJ, TYK, LS, JEN, and GB conceived and designed the research. GB, AA, UR, TJ, AF, CV, HH and BR performed testing. GB, AA, CMTH, AF, UR, JEN, TJ, JG, and BR analyzed data and interpreted results. BJB, CM, TAI and HH interpreted the results and edited the discussion. GB and AA prepared figures and drafted the manuscript. All authors edited, revised and approved the final version of the manuscript submitted for publication

## Funding

This study was supported by the National Institute of Diabetes and Digestive and Kidney Diseases grants R03 DK114502 (to BR), R01DK125794 (to JG), and R01DK129793 (to BR), TL1DK139565 (to CH and AA). Support was also provided by Dialysis Clinics grant C-4112 (to BR).

## Acknowledgments

We thank all the study participants for being part of this trial. Thank you to all exercise trainers involved in this study from California State University of Sacramento and Fullerton including Morgan Andrew, Brittany Lyon, Stephen Gonzales, Catherine Welsh Osuna, and Maggie Fray. We could not have conducted this study without the support of Erica Goude and her team of research kinesiologists and coordinators at the Neuromuscular Research Laboratory (NMRL) at UC Davis Health, particularly Alina Nicorici, Lynea Kaethler, and Amanda Lopez. We thank the fantastic junior specialists Radhika Batra, Harshanna Badhesha, and Geraldine Portillo for their help with study coordination, data entry and collection. Finally, we thank David Bendahan, Gerald Sonico, and Yann Le Fur, and Eric Shankland for their expert advice in setting up the MRS experiment as well as Amy Vansdadia, Julianna Porter, Lauren Andrews, and Katelyn Nielsen for their help in processing the MRS data.

---

**Supplemental Figure 1.**
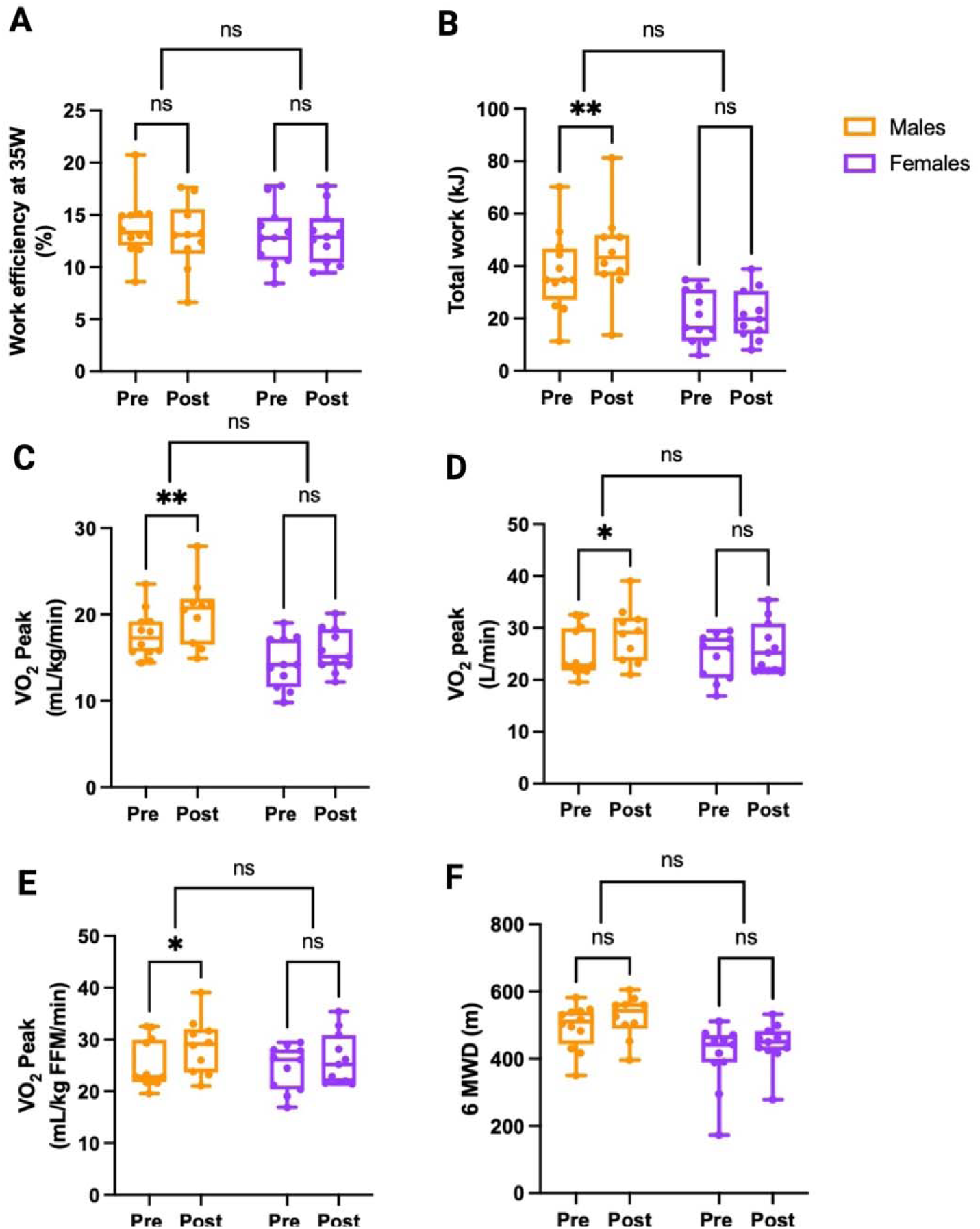
Changes in physical performance, cardiorespiratory fitness, and efficiency among males (n=12) and females (n=11) randomized to exercise.

**Supplemental Figure 2.**
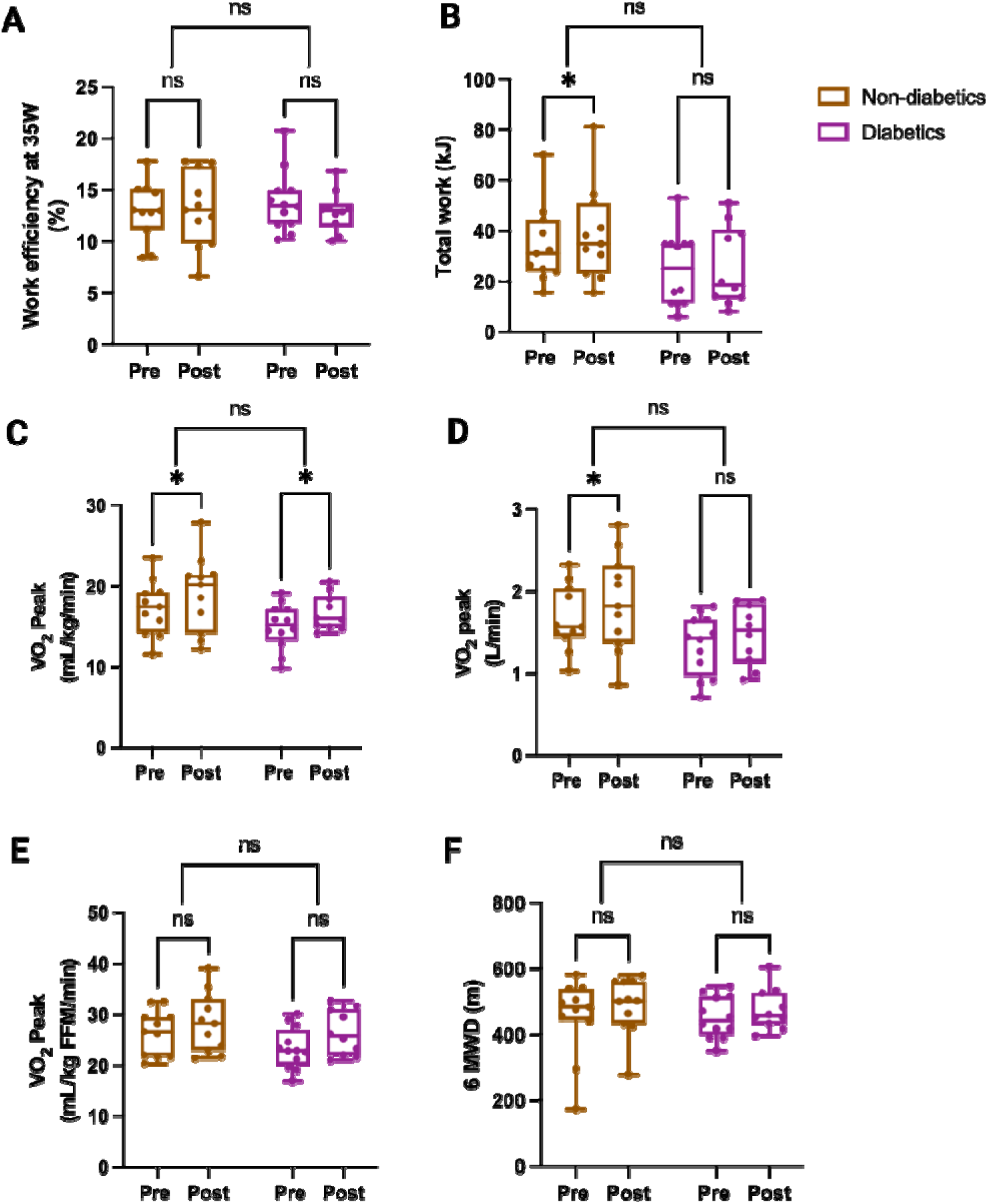
Changes in physical performance and cardiorespiratory fitnes among participants with (n=12) and without (n=11) diabetes randomized to exercise.

**Supplemental Table 1.** Full list of study exclusion criteria.

| **Kidney function Demographics** | **Contraindication to participate in vigorous exercise or MRI** |
| --- | --- |
| Estimated glomerular filtration rate (eGFR) 60ml/min per 1.73m<sup>2</sup> or greater | Pacemaker |
| Age <30 or >75 years old | History of severe heart disease/disorders: coronary artery bypass graft (CABG) surgery, atrial fibrillation |
| Weight >135 kg (limit of MRI table) | Abnormal physiological and cardiac responses (i.e., coronary ischemia) to cardiopulmonary testing (CPET) |
| Not sedentary defined as self-reporting more than 1 day/week of structured endurance exercise or resistance exercise lasting more than 60min in the past year.<br>People bicycling as a mode of transportation to/from work > 1 day/week. | Wheelchair dependence or other disability that precludes physical exercise |
| Non-English speaking or inability to provide consent without a proxy respondent. | Oxygen dependent chronic obstructive pulmonary disease (COPD) |
| **Medical Conditions** | **Medications** |
| Uncontrolled diabetes with a HgbA1c >8.5 | Drugs that alter mitochondrial function: muscle relaxants, oral steroids, anti-viral medications, oral calcineurin inhibitors, antiepileptics, antipsychotics. |
| Uncontrolled hypertension defined as systolic blood pressure >170mmHg or diastolic blood pressure >100mmHg | Supplements known to alter mitochondrial function: coenzyme Q10, nicotinamide riboside, antioxidants. |
| Anemia (Hgb <9g/dL) | Anticoagulant use or dual antiplatelet therapy |
| Current or previous transplantation or on chronic dialysis or expectation to start dialysis in 6 months. | **Additional Criteria** |
| Current pregnancy | Current participation in an interventional trial |
| HIV infection or hepatitis viral infection | Implants that prohibit MRI measurements or trauma involving metal fragments |
| Decompensated cirrhosis | History of clotting disorder (Deep venous thrombosis, pulmonary embolism) or bleeding disorder. |
| Active malignant cancer other than non-melanomatous skin cancer | Current substance abuse |
| Vascular stent: bare metal or any recently placed (within 6 months) | Institutionalization |

**Supplemental Table 2.** Adverse events in each group. Treatment emergent adverse events were counted from the start of randomization until the end of the post-treatment visit. *Patient required cardiac catheterization and drug eluting stent placement for left anterior descending lesion.

| **Adverse Event** | **Usual care** | **Exercise** |
| --- | --- | --- |
| Hypertensive urgency | 0 (0%) | 1 (4%) |
| Hypotension requiring BP medication reduction | 0 (0%) | 1 (4%) |
| PD catheter insertion (not used) | 0 (0%) | 1 (4%) |
| Progression of renal disease requiring initiation of PD | 1(11%) | 0 (0%) |
| Chest pain with intercourse | 0 (0%) | 1 (4%) |
| Tachycardia and dyspnea during CPET* | 1 (11%) | 0 (0%) |

**Supplemental Table 3.** Physical performance and metabolic efficiency endpoints comparing usual care and exercise at baseline and after the 12-week intervention (n=32). Linear mixed effects modeling was used to estimate the effect of exercise compared to usual care. Mean differences, 95% CIs, and *P*-values are shown. The values in bold represent *P* <0.05.

| Endpoint | Usual Care |  | Exercise |  | Within group differences |  | Between group differences |  |
| --- | --- | --- | --- | --- | --- | --- | --- | --- |
|  | Baseline mean (SD) | 12 weeks mean (SD) | Baseline mean (SD) | 12 weeks mean (SD) | Usual care Beta (95% CI) | Exercise Beta (95% CI) | Exercise-usual care Beta (95% CI) | Treatment *P*-value |
| **Graded Exercise Test** |  |  |  |  |  |  |  |  |
| Total Test Duration | 386 (168) | 423 (127) | 341 (161) | 457 (136) | -17 (-41, 7) | **39 (22, 56)** | **57 (27, 87)** | <0.001 |
| Peak Work Rate (W) | 101.7 (41.5) | 95.7 (37.8) | 104.6 (29.1) | 112.4 (32.5) | -0.38 (-10.9, 10.2) | **8.25 (1.24, 15.3)** | 8.85 (-3.82, 21.5) | 0.20 |
| Peak RER | 0.92 (0.23) | 1.02 (0.21) | 0.93 (0.20) | 1.04 (0.28) | 0.03 (-0.09, 0.15) | 0.01 (-0.07, 0.09) | -0.03 (-0.17, 0.12) | 0.69 |
| Peak RPE | 6.9 (2.7) | 7.9 (1.9) | 7.6 (2.5) | 8.3 (1.5) | 0.81 (-0.35, 1.99) | 0.50 (-0.50, 1.51) | -0.45 (-1.95, 1.04) | 0.56 |
| VO<sub>2</sub> at 35W | 0.92 (0.23) | 1.02 (0.21) | 0.93 (0.20) | 1.04 (0.28) | 0.03 (-0.09, 0.15) | 0.01 (-0.07, 0.09) | -0.03 (-0.17, 0.12) | 0.69 |
| RER at 35W | 0.75 (0.12) | 0.81 (0.18) | 0.76 (0.13) | 0.76 (0.13) | 0.03 (-0.05, 0.12) | -0.001 (-0.03, 0.027) | -0.037 (-0.129, 0.054) | 0.44 |
| **6-minute Walk Test** |  |  |  |  |  |  |  |  |
| Walking Economy (mlO<sub>2</sub>/m) | 0.18 (0.02) | 0.19 (0.03) | 0.19 (0.02) | 0.19 (0.02) | 0.016 (0.001, 0.032) | 0.009 (-0.0008, 0.019) | -0.007 (-0.026, 0.011) | 0.41 |

## Notes

### Competing Interest Statement

The authors have declared no competing interest.

### Clinical Trial

NCT 02923063

### Funding Statement

This study was supported by the National Institute of Diabetes and Digestive and Kidney Diseasesvgrants R03 DK114502, R01DK125794, and R01DK129793 , TL1DK139565. Support was also provided by Dialysis Clinics grant C-4112.

### Author Declarations

Ethics committee/IRB of University of California Davis gave ethical approval of this work (ID 1343904-21)

## References

1. Chronic kidney disease in the United States, 2021. Pamphlet (or booklet). 2021;

2. Roshanravan B, Patel KV, Fried LF, et al. Association of Muscle Endurance, Fatigability, and Strength With Functional Limitation and Mortality in the Health Aging and Body Composition Study. J Gerontol A Biol Sci Med Sci. Feb 2017;72(2):284–291. doi:10.1093/gerona/glw210

3. Roshanravan B, Robinson-Cohen C, Patel KV, et al. Association between physical performance and all-cause mortality in CKD. J Am Soc Nephrol. Apr 2013;24(5):822–30. doi:10.1681/ASN.2012070702

4. Jhamb M, Argyropoulos C, Steel JL, et al. Correlates and outcomes of fatigue among incident dialysis patients. Clin J Am Soc Nephrol. Nov 2009;4(11):1779–86. doi:10.2215/CJN.00190109

5. Bossola M, Di Stasio E, Antocicco M, Panico L, Pepe G, Tazza L. Fatigue Is Associated with Increased Risk of Mortality in Patients on Chronic Hemodialysis. Nephron. 2015;130(2):113–8. doi:10.1159/000430827

6. Kurella Tamura M, Covinsky KE, Chertow GM, Yaffe K, Landefeld CS, McCulloch CE. Functional status of elderly adults before and after initiation of dialysis. N Engl J Med. Oct 15 2009;361(16):1539–47. doi:10.1056/NEJMoa0904655

7. Roshanravan B, Patel KV. Assessment of physical functioning in the clinical care of the patient with advanced kidney disease. Semin Dial. Jul 2019;32(4):351–360. doi:10.1111/sdi.12813

8. Tamaki M, Miyashita K, Wakino S, Mitsuishi M, Hayashi K, Itoh H. Chronic kidney disease reduces muscle mitochondria and exercise endurance and its exacerbation by dietary protein through inactivation of pyruvate dehydrogenase. Kidney Int. Jun 2014;85(6):1330–9. doi:10.1038/ki.2013.473

9. Tian Q, Mitchell BA, Zampino M, Fishbein KW, Spencer RG, Ferrucci L. Muscle mitochondrial energetics predicts mobility decline in well-functioning older adults: The baltimore longitudinal study of aging. Aging cell. Feb 2022;21(2):e13552. doi:10.1111/acel.13552

10. Santanasto AJ, Glynn NW, Jubrias SA, et al. Skeletal Muscle Mitochondrial Function and Fatigability in Older Adults. J Gerontol A Biol Sci Med Sci. Nov 2015;70(11):1379–85. doi:10.1093/gerona/glu134

11. Roshanravan B, Kestenbaum B, Gamboa J, et al. CKD and Muscle Mitochondrial Energetics. American Journal of Kidney Diseases. 2016/10/01/ 2016;68(4):658-659. 10.1053/j.ajkd.2016.05.011

12. Kestenbaum B, Gamboa J, Liu S, et al. Impaired skeletal muscle mitochondrial bioenergetics and physical performance in chronic kidney disease. JCI Insight. Mar 12 2020;5(5)doi:10.1172/jci.insight.133289

13. Gamboa JL, Roshanravan B, Towse T, et al. Skeletal Muscle Mitochondrial Dysfunction Is Present in Patients with CKD before Initiation of Maintenance Hemodialysis. Clin J Am Soc Nephrol. Jul 1 2020;15(7):926–936. doi:10.2215/CJN.10320819

14. Roshanravan B, Gamboa J, Wilund K. Exercise and CKD: Skeletal Muscle Dysfunction and Practical Application of Exercise to Prevent and Treat Physical Impairments in CKD. Am J Kidney Dis. Jun 2017;69(6):837–852. doi:10.1053/j.ajkd.2017.01.051

15. Hayden CMT, Begue G, Gamboa JL, Baar K, Roshanravan B. Review of Exercise Interventions to Improve Clinical Outcomes in Nondialysis CKD. Kidney International Reports. 2024/08/02/ 2024;10.1016/j.ekir.2024.07.032

16. Bernier-Jean A, Beruni NA, Bondonno NP, et al. Exercise training for adults undergoing maintenance dialysis. Cochrane Database Syst Rev. Jan 12 2022;1(1):CD014653. doi:10.1002/14651858.CD014653

17. Baiao VM, Cunha VA, Duarte MP, et al. Effects of Exercise on Inflammatory Markers in Individuals with Chronic Kidney Disease: A Systematic Review and Meta-Analysis. Metabolites. Jun 27 2023;13(7)doi:10.3390/metabo13070795

18. Wu X, Yang L, Wang Y, Wang C, Hu R, Wu Y. Effects of combined aerobic and resistance exercise on renal function in adult patients with chronic kidney disease: a systematic review and meta-analysis. Clinical Rehabilitation. 2020/07/01 2020;34(7):851-865. doi:10.1177/0269215520924459

19. MoTr PACSG, Jakicic JM, Kohrt WM, et al. Molecular Transducers of Physical Activity Consortium (MoTrPAC): human studies design and protocol. J Appl Physiol (1985). Sep 1 2024;137(3):473-493. doi:10.1152/japplphysiol.00102.2024

20. Gamborg M, Hvid LG, Thrue C, et al. Muscle Strength and Power in People With Parkinson Disease: A Systematic Review and Meta-analysis. J Neurol Phys Ther. Jan 1 2023;47(1):3–15. doi:10.1097/NPT.0000000000000421

21. Jacobsen SC, Brons C, Bork-Jensen J, et al. Effects of short-term high-fat overfeeding on genome-wide DNA methylation in the skeletal muscle of healthy young men. Diabetologia. Dec 2012;55(12):3341–9. doi:10.1007/s00125-012-2717-8

22. Aging NIo. Workout to Go. October 2011

23. NIDDK. Eating Right for Chronic Kidney Disease. 2016;

24. Coyle EF, Sidossis LS, Horowitz JF, Beltz JD. Cycling efficiency is related to the percentage of type I muscle fibers. Med Sci Sports Exerc. Jul 1992;24(7):782–8.

25. Matomäki P, Linnamo V, Kyröläinen H. A Comparison of Methodological Approaches to Measuring Cycling Mechanical Efficiency. Sports Med Open. Jun 10 2019;5(1):23. doi:10.1186/s40798-019-0196-x

26. Rehman U, Begue G, Ahmadi A, et al. An ergometer dataset to measure muscle bioenergetics with magnetic resonance techniques. Data Brief. Dec 2024;57:111114. doi:10.1016/j.dib.2024.111114

27. Kaminsky LA, Arena R, Myers J, et al. Updated Reference Standards for Cardiorespiratory Fitness Measured with Cardiopulmonary Exercise Testing: Data from the Fitness Registry and the Importance of Exercise National Database (FRIEND). Mayo Clin Proc. Feb 2022;97(2):285–293. doi:10.1016/j.mayocp.2021.08.020

28. Zwisler AD, Norton RJ, Dean SG, et al. Home-based cardiac rehabilitation for people with heart failure: A systematic review and meta-analysis. Int J Cardiol. Oct 15 2016;221:963–9. doi:10.1016/j.ijcard.2016.06.207

29. Gonzalez-Freire M, Scalzo P, D’Agostino J, et al. Skeletal muscle ex vivo mitochondrial respiration parallels decline in vivo oxidative capacity, cardiorespiratory fitness, and muscle strength: The Baltimore Longitudinal Study of Aging. Aging cell. Apr 2018;17(2)doi:10.1111/acel.12725

30. McCully KK, Fielding RA, Evans WJ, Leigh JS, Jr., Posner JD. Relationships between in vivo and in vitro measurements of metabolism in young and old human calf muscles. J Appl Physiol (1985). Aug 1993;75(2):813-9. doi:10.1152/jappl.1993.75.2.813

31. Mau T, Lui LY, Distefano G, et al. Mitochondrial Energetics in Skeletal Muscle Are Associated With Leg Power and Cardiorespiratory Fitness in the Study of Muscle, Mobility and Aging. J Gerontol A Biol Sci Med Sci. Aug 2 2023;78(8):1367–1375. doi:10.1093/gerona/glac238

32. Bohannon RW, Crouch R. Minimal clinically important difference for change in 6-minute walk test distance of adults with pathology: a systematic review. J Eval Clin Pract. Apr 2017;23(2):377–381. doi:10.1111/jep.12629

33. Sietsema KE, Amato A, Adler SG, Brass EP. Exercise capacity as a predictor of survival among ambulatory patients with end-stage renal disease. Kidney Int. Feb 2004;65(2):719–24. doi:10.1111/j.1523-1755.2004.00411.x

34. Kokkinos P, Faselis C, Samuel IBH, et al. Cardiorespiratory Fitness and Mortality Risk Across the Spectra of Age, Race, and Sex. J Am Coll Cardiol. Aug 9 2022;80(6):598–609. doi:10.1016/j.jacc.2022.05.031

35. Mahatme S, K V, Kumar N, Rao V, Kovela RK, Sinha MK. Impact of high-intensity interval training on cardio-metabolic health outcomes and mitochondrial function in older adults: a review. Med Pharm Rep. Apr 2022;95(2):115–130. doi:10.15386/mpr-2201

36. Milanovic Z, Sporis G, Weston M. Effectiveness of High-Intensity Interval Training (HIT) and Continuous Endurance Training for VO2max Improvements: A Systematic Review and Meta-Analysis of Controlled Trials. Sports Med. Oct 2015;45(10):1469–81. doi:10.1007/s40279-015-0365-0

37. Macdonald JH, Marcora SM, Jibani M, Phanish MK, Holly J, Lemmey AB. Intradialytic exercise as anabolic therapy in haemodialysis patients -- a pilot study. Clin Physiol Funct Imaging. Mar 2005;25(2):113–8. doi:10.1111/j.1475-097X.2004.00600.x

38. Cachofeiro V, Goicochea M, de Vinuesa SG, Oubiña P, Lahera V, Luño J. Oxidative stress and inflammation, a link between chronic kidney disease and cardiovascular disease: New strategies to prevent cardiovascular risk in chronic kidney disease. Kidney International. 2008/12/01/ 2008;74:S4-S9. 10.1038/ki.2008.516

39. Cobo G, Lindholm B, Stenvinkel P. Chronic inflammation in end-stage renal disease and dialysis. Nephrol Dial Transplant. Oct 1 2018;33(suppl_3):iii35-iii40. doi:10.1093/ndt/gfy175

40. Wu L, Liu Y, Wu L, Yang J, Jiang T, Li M. Effects of exercise on markers of inflammation and indicators of nutrition in patients with chronic kidney disease: a systematic review and meta-analysis. Int Urol Nephrol. Apr 2022;54(4):815–826. doi:10.1007/s11255-021-02949-w

41. Bickel M. The role of interleukin-8 in inflammation and mechanisms of regulation. J Periodontol. May 1993;64(5 Suppl):456-60.

42. Wang T. Searching for the link between inflammaging and sarcopenia. Ageing Res Rev. May 2022;77:101611. doi:10.1016/j.arr.2022.101611

43. Pan L, Xie W, Fu X, et al. Inflammation and sarcopenia: A focus on circulating inflammatory cytokines. Exp Gerontol. Oct 15 2021;154:111544. doi:10.1016/j.exger.2021.111544

44. Beavers KM, Hsu FC, Isom S, et al. Long-term physical activity and inflammatory biomarkers in older adults. Med Sci Sports Exerc. Dec 2010;42(12):2189–96. doi:10.1249/MSS.0b013e3181e3ac80

45. Petersen AM, Pedersen BK. The anti-inflammatory effect of exercise. J Appl Physiol (1985). Apr 2005;98(4):1154-62. doi:10.1152/japplphysiol.00164.2004

46. Alizaei Yousefabadi H, Niyazi A, Alaee S, Fathi M, Mohammad Rahimi GR. Anti-Inflammatory Effects of Exercise on Metabolic Syndrome Patients: A Systematic Review and Meta-Analysis. Biol Res Nurs. Apr 2021;23(2):280–292. doi:10.1177/1099800420958068

47. Kojima S, Usui N, Uehata A, et al. Relationships between frailty and exercise capacity in patients undergoing hemodialysis: A cross-sectional study. Geriatr Gerontol Int. Nov 2023;23(11):795–802. doi:10.1111/ggi.14681

