## Supplemental Methods for "A Pilot Randomized Trial of Home-Based, Video-Supervised Exercise on Muscle Metabolism and Physical Endurance in Chronic Kidney Disease"

*6-minute walk test (6MWT)*

Participants were instructed to walk back and forth around cones on a 25m hallway as fast as possible for 6min and the total distance covered was measured (6MWD). Oxygen consumption (VO_2_) was collected as 30s averages using a breath-by-breath metabolic analyzer system (Cosmed K5) and VO_2_ data points were averaged over the last 3min (steady-state VO_2_) to calculate walking economy as relative VO_2_ (ml/kg/min) divided by walking speed (m/min).

*Graded cycle exercise testing (GXT)*

Oxygen consumption (VO_2_) and carbon dioxide production (VCO_2_) were measured as 30s averages at baseline and during the GXT using a breath-by-breath metabolic analyzer system (Cosmed K5). Fuel utilization was assessed using the respiratory exchange ratio (i.e., RER, VCO_2_/VO_2_). After resting for 3min on the cycle ergometer to collect baseline data, participants conducted a warm-up at 10W for 3min. Then, the load was increased by 25W every 2min until participants reached exhaustion and proceeded to a cool-down for 5min at 10W. Peak VO_2_, RER, and RPE were obtained during the final stage of the GXT (before cool-down). During the test, participants were asked to maintain a cadence of 50-70RPM. Manual blood pressures and RPEs using the modified Borg scale (scale of 1-10) were taken at baseline, during GXT and cool-down. Cardiac rhythm was monitored during the test using a 12-leads electrocardiogram (Quark C12x, Cosmed). Abnormal physiological and cardiac responses during the GXT would lead to premature test termination and would exclude participants from continuing into the study (**Supplemental Table 1**).

*In-vivo muscle oxidative capacity (^31^P-MRS)*

Participants were positioned supine and feet-first on the spectrometer bed. A half-cylinder foam was placed under the right knee joint to create about a 40° bend referenced to the neutral position aligning the greater trochanter with the lateral malleolus. A 12cm PulseTeq ^31^P/^1^H surface coil (PulseTeq, Chobham, Woking, United Kingdom) was secured over the quadriceps (about halfway between the knee and thigh joint). Upon receiving the start signal, participants began to kick freely for 42s at 1Hz guided by a metronome. For the next 6mins after exercise cessation, exercise recovery signal was recorded. JMRUI version 6.0 (MRUI Consortium, European Union) processed the spectra with an exponential line broadening of 10 Hz. ^31^P-MRS assessment and k_PCr_ calculations were conducted as previously described^24^.

*Body Composition and Resting Energy Expenditure*

Body composition and resting energy expenditure (REE) were measured pre- and post-intervention in the morning after an 8-12hr fast. Participants were instructed to refrain from strenuous exercise, caffeine and alcohol as well as remain hydrated 24hr before testing. After participants rested in a supine position for 3-5min, body composition was assessed using bioimpedance spectroscopy (Impedimed SFB7). Then, REE was measured in a seated position using a facemask and breath-by-breath metabolic analyzer system (Cosmed K5). Data were collected as 1min averages for 15min. REE in kcal/day was computed with the last 10min of data collection using the Weir equation (Omnia software 2.1.1.0).

*Plasma Cytokines*

Fasted blood draws were collected in 10ml *EDTA*-tubes and centrifuged at 2,000g for 10min at room temperature. Plasma was collected from the top layer, aliquoted and stored in cryovials at -80°C until analysis. Cytokines concentration (pg/ml) was computed from the average duplicates according to standard curves. The mean coefficient of variation was 3.16 ±5.5% across all measured cytokines.
